# Instrumental Activities of Daily Living in Older Adults with Epilepsy: A Cross-Sectional and Longitudinal Multicenter Study

**DOI:** 10.64898/2026.06.13.26355582

**Authors:** Ifrah Zawar, Anny Reyes, Kayela Arrotta, Bruce P. Hermann, Robyn M. Busch, Mark Quigg, Jaideep Kapur, Aaron F. Struck, Vineet Punia, Alena Stasenko, McKenna E. Williams, Katherine J. Bangen, Irene Wang, Jerry J. Shih, Lisa Ferguson, Jana E. Jones, Carrie R. McDonald

**Author notes:** Corresponding Author: Ifrah Zawar, MD, MS-CR, POB 800994, HSC, Department of Neurology, University of Virginia, School of Medicine, Charlottesville, VA 22908.

## Abstract

**Objective:** Instrumental activities of daily living (IADLs) represent a critical but understudied measure of day-to-day function in persons with epilepsy(PWE). In the multicenter Brain Aging and Cognition in Epilepsy (BrACE) study of PWE aged ≥55 years, we examined the proportion, clinical correlates, epilepsy-related predictors, and longitudinal trajectory of IADL impairment.

**Methods:** IADLs were assessed using the Functional Activities Questionnaire(FAQ; range=0–30; higher=more impaired); FAQ≥2 defines MCI-level impairment and FAQ≥5 dementia-level functional impairment. Multivariable logistic regression identified predictors of baseline function. Global cognition (Montreal Cognitive Assessment [MoCA]), individual cognitive measures, and quality of life (QOL) were compared between the impaired and unimpaired groups. Linear regression evaluated predictors of longitudinal functional decline.

**Results:** Of 57 participants(mean age=66.6±7.2 years; female=52.6%), 38.6%(n=22) had MCI-level functional impairment and 17.5%(n=10) had dementia-level functional impairment. In univariate analyses, worse FAQ scores were associated with lower education, higher area deprivation index, early-onset epilepsy (EOE < 60 years), antiseizure medication polytherapy, and epilepsy localization (all p < 0.05). In multivariable analysis, temporal lobe epilepsy (OR=4.46, 95%-CI=1.09–21.83,p=0.047), EOE(OR=7.14,95%-CI=1.16–59.97,p=0.046), and lower education(OR=0.70,95%-CI=0.49–0.93,p=0.025) remained independently associated with baseline MCI-level functional-impairment. Lower education (OR=0.55,95%-CI=0.29-0.84, p=0.021) was the only factor associated with dementia-level IADL-impairment. IADL-impaired participants demonstrated lower verbal memory scores(adjusted-p=0.041) and MoCA(adjusted-p<0.001), particularly in visuospatial/executive function, attention, and memory subscores, and worse QOL(adjusted-p=0.041). IADL-impairment was greatest in financially-mediated and memory-dependent tasks.

Longitudinally, EOE(β=7.51,95%-CI=1.92–13.10,p=0.017) and older age(β=0.38,95%-CI=0.12–0.65,p=0.012) predicted greater functional decline. Nearly one-third progressed to significantly worse function over a two-year period, with 15.4% progressing from MCI-level to dementia-level impairment and 15.4% from normal function to MCI-level IADL-impairment.

**Conclusions:** Functional impairment affects ∼40% of older PWE, with ∼1-in-6 experiencing functional impairment comparable to overt dementias. Temporal lobe localization, EOE, lower education, and poorer cognition are important determinants of baseline functional status. EOE and older age predict accelerated functional decline, suggesting that cumulative disease burden and aging-related processes may drive functional deterioration. These findings provide one of the first epilepsy-specific longitudinal characterizations of IADL impairment and support routine functional assessment in PWE.

**Key Points:**

- Functional impairment is common in older PWE, affecting **∼**40%, with 1 in 6 showing dementia-level functional impairment.
- Temporal lobe epilepsy localization, early-onset epilepsy, and lower education are independently associated with baseline MCI-level functional impairment.
- Lower education was the only independent predictor of dementia-level IADL impairment.
- IADL impairment was associated with worse verbal memory, global cognition, and quality of life.
- Over two years, early-onset epilepsy and older age predicted greater functional decline, with nearly one-third progressing to worse functional status.

## Introduction

Epilepsy is the third most common neurological disorder in older adults, affecting over 700,000 Medicare beneficiaries in the United States, with late-onset epilepsy (onset ≥60 years) comprising the fastest-growing subgroup of individuals with epilepsy.^1^ Beyond seizures, cognitive comorbidities are well-documented in these individuals. Approximately two-thirds of older people with epilepsy (PWE) exhibit some degree of cognitive impairment, a rate comparable to that observed in mild cognitive impairment (MCI).^2,3^ The presence of comorbid epilepsy in persons with Alzheimer’s disease and related dementias (ADRD) is associated with faster cognitive decline, worse judgment, and premature mortality.^4–8^ Despite the high prevalence of cognitive dysfunction among older PWE, its impact on day-to-day functioning remains understudied.

Instrumental activities of daily living (IADLs), encompassing complex real-world tasks such as financial management, medication adherence, meal preparation, and travel, represent a critical interface between cognitive function and independent living.^9^ The downstream functional consequences of epilepsy on IADLs remain substantially under-characterized in epilepsy-specific cohorts. The Pfeffer Functional Activities Questionnaire (FAQ) is a validated, informant-based instrument widely used in dementia research to quantify IADL performance, with a high discriminative ability between cognitive groups.^10,11^ FAQ is used to assess and track the severity of functional dependence.^11^

Our recent study motivates the present investigation. We demonstrated in a large Alzheimer’s Disease Research Center (ADRC) cohort (N = 50,724) that active comorbid epilepsy independently predicts worse baseline FAQ and accelerated longitudinal FAQ decline, with taxes, travel, bills, and appointment management most affected.^9^ However, the ADRC data lacked epilepsy-specific characterization, including seizure localization, timing of epilepsy onset, types of seizures, drug-resistant epilepsy (DRE) status, and detailed neuropsychological profiling, afforded by a prospectively enrolled epilepsy cohort.

In this multicenter well-characterized epilepsy cohort, this study aimed to fill this knowledge gap by: (1) characterizing the proportion and profile of FAQ-defined functional impairment in older PWE; (2) identifying epilepsy-specific and demographic predictors of functional impairment; (3) examining associations between functional impairment and both global cognition and individual cognitive measures, and quality of life; and finally (4) exploring baseline predictors of longitudinal FAQ trajectory in older PWE.

## Methods

### Study Design and Participants

Brain Aging and Cognition in Epilepsy (BrACE) is a prospective, multicenter study that includes an observational cohort of older adults with early-onset epilepsy (EOE) and late-onset unexplained epilepsy (LOUE) with no identifiable cause (e.g., stroke, tumor) enrolled at three tertiary academic epilepsy centers (University of California, San Diego, Cleveland Clinic, and University of Wisconsin-Madison). BrACE participants undergo comprehensive demographic, epilepsy, neuropsychological, and functional assessment at enrollment and longitudinal follow-up visits, enabling an epilepsy-specific examination of IADL determinants.

### Inclusion and Exclusion Criteria

Eligible participants met the following BrACE inclusion criteria: (1) age ≥55 years, (2) confirmed diagnosis of focal epilepsy by a board-certified epilepsy specialist, (3) no prior epilepsy-related neurosurgery, stroke, brain tumor, or other space-occupying lesions, and (4) sufficient English fluency to complete a neuropsychological assessment. Those with overt dementia at study enrollment were excluded. The present analysis was additionally restricted to participants with FAQ data completed by a study partner at the baseline visit. All participants and study partners provided written informed consent.

### Clinical, Demographic, and Epilepsy Measures

Demographic, clinical, and epilepsy-related characteristics were collected through patient questionnaires and electronic medical record review. Variables included age, sex, years of education, area deprivation index^12^ (ADI; state decile and national percentile), number of antiseizure medications (ASMs), whether participants had drug-resistant (DRE) or drug-responsive epilepsy, active epilepsy duration, seizure localization (frontal, temporal, or other), and age of epilepsy onset. Participants were classified as EOE (seizure onset <60 years) or LOUE (seizure onset ≥60 years).

### Functional Assessment

The Pfeffer FAQ is a validated, 10-item informant-based measure of IADLs assessing financial management, tax preparation, shopping, games/hobbies, appliance use, meal preparation, event tracking, attention to media, appointment and medication management, and travel.^13^ Items are scored 0 (normal) to 3 (dependent), with a total range of 0–30; higher scores indicate greater IADL impairment.

Recent large validation studies suggest that FAQ ≥2 can distinguish those with normal cognitive function from those with MCI, and that FAQ >5 defines the functional impairment cut-off for dementia.^13^ Given that people with dementia were excluded from BrACE at baseline, FAQ ≥2 was used to define functional impairment for the primary outcome.^9^ In sensitivity analysis, FAQ ≥5 was also considered. Study partners completed the FAQ at enrollment and the two-year follow-up.

### Neuropsychological and Quality-of-Life Measures

All participants underwent a comprehensive neuropsychological battery assessing global cognitive and individual measures of cognition, including memory, language, executive function, and attention/processing speed.

Global cognition was measured with the Montreal Cognitive Assessment (MoCA; total and domain subscores).^14^ MoCA domain subscores further analyzed included Naming, Attention, Abstraction, Memory/Delayed Recall, and Visuospatial/Executive function.

Memory was assessed with the Wechsler Memory Scale (WMS-IV) Logical Memory II delayed recall and RAVLT delayed recall, and learning over trials. Language was assessed with Animal Naming and the Multilingual Naming Test(MINT).^15^ Executive function was assessed using the Letter Fluency and Trail-Making Test, Part B.^16^ Processing speed and attention were evaluated with the Trail-Making Test Part A. All measures were scored using demographically corrected norms and then converted to standard scores (T-scores).

The Alzheimer’s Disease Assessment Scale-Cognitive Subscale (ADAS-Cog; 13-item total, range 0–85, higher scores=greater cognitive impairment) provided an additional objective cognitive index, with particular sensitivity to prominent symptoms of dementia.^17^ It yields a global cognitive severity score used in Alzheimer’s disease (AD) and MCI clinical trials.^17^

Subjective cognitive function was quantified with the Everyday Cognition (ECog) scale (informant-rated; higher scores indicate worse subjective cognition).^18^

Quality of life was assessed with the Quality of Life in Epilepsy Inventory (QOLIE-31; overall T-score; higher scores indicate better quality of life).^19^

### Statistical Analyses

#### Baseline Characteristics and Instrumental Activities of Daily Living

Baseline characteristics were summarized using the mean and standard deviation for continuous variables and proportions and percentages for categorical variables. Univariate associations between baseline characteristics and the FAQ total score were evaluated using Spearman’s correlation for continuous predictors, Wilcoxon rank-sum tests for binary categorical predictors, and the Kruskal-Wallis test for three-group categorical predictors (≥3 levels), given the skewness of the FAQ distribution (**Figure 1**).

**Figure 1:**
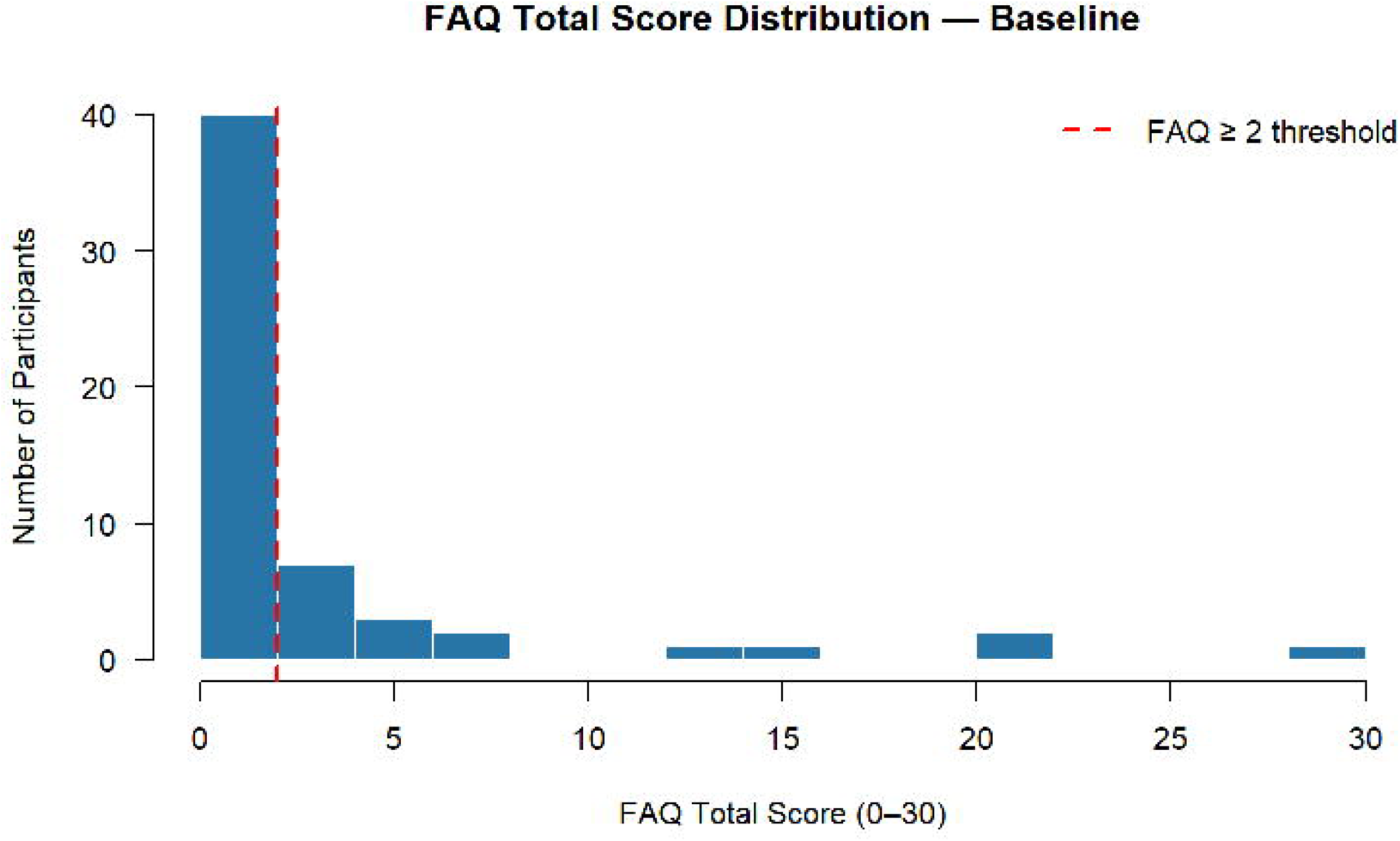
Functional Activities Questionnaire (FAQ) Total Distribution at Baseline.

#### Predictors of Baseline Functional Impairment

Multivariable logistic regression with FAQ ≥2 as the binary outcome was used to identify independent predictors of functional impairment comparable to MCI. FAQ was treated as a categorical dependent variable, and the following independent variables were considered: age, number of ASMs, epilepsy onset group (LOUE versus EOE), seizure localization (temporal versus frontal/other; other [N=4] collapsed with frontal), years of education and ADI decile. In sensitivity analysis, logistic regression with FAQ ≥5 as the binary outcome was used to identify independent predictors of functional impairment comparable to dementia.

#### Functional Impairment and Cognition

To examine neuropsychological and quality-of-life correlates of functional impairment, independent samples t-tests compared mean scores across 21 neuropsychological and quality-of-life measures between impaired (FAQ ≥2) and unimpaired (FAQ < 2) groups; all p-values were corrected using the Benjamini-Hochberg false discovery rate method.

#### Individual Instrumental Activities of Daily Living

In an exploratory analysis, differences in individual FAQ item scores between EOE versus LOUE groups, DRE versus non-DRE groups, and temporal versus non-temporal localization (Frontal, others) groups were assessed using Wilcoxon rank-sum tests.

#### Predictors of the Longitudinal Trajectory of Instrumental Activities of Daily Living

For the exploratory longitudinal analysis, a change-score approach was used, given the small sample size (N = 13 participants with FAQ at two visits 2 years apart). The FAQ change was calculated as follows: follow-up minus baseline FAQ total. In addition to continuous FAQ change scores, longitudinal transitions across functional impairment categories were examined using established FAQ thresholds: no functional impairment (FAQ <2), MCI-level functional impairment (FAQ ≥2), and dementia-level functional impairment (FAQ >5). Clinically meaningful decline was defined as progression to a more severe functional impairment category at follow-up. Baseline predictors of FAQ longitudinal change were examined using multivariable linear regression models.

Missing data were excluded from the analyses because of the small sample size. Statistical analyses were performed using R version 4.4.2. Statistical significance was defined as two-sided α = 0.05.

## Results

### Baseline Characteristics

Baseline visit FAQ data were available for 57 participants (30 female [52.6%]; mean age 66.6 ± 7.2 years) (**Table 1**). The mean FAQ total score was 3.0 ± 6.0. Twenty-two participants (38.6%) met the threshold for functional impairment comparable to MCI (FAQ ≥2) [**Figure 1**], and 10 (17.5%) met the threshold for functional impairment comparable to dementia. The average years of education was 14.9 ± 2.7. The majority of participants were White (93.0%). The mean ADI national percentile was 52.1 ± 24.6.

**Table 1.** Baseline Cohort Characteristics and Associations with Functional Activities Questionnaire (FAQ) Total Score

| Table 1: Characteristics | Total Study Sample (N = 57) | FAQ Scores (Average ± SD) | p-value |
| --- | --- | --- | --- |
| FAQ Total Score | N = 57 | 3.0 ± 6.0 | - |
| Number impaired (FAQ ≥ 2), N (%) | 22 (38.6%) | - | - |
| **Demographics** |  |  |  |
| Age in years, Mean ± SD | 66.6 ± 7.2 | Rho=-0.175 | 0.194^ |
| Sex — Male, N (%) | 27 (47.4%) | 3.1 ± 6.2 | 0.716# |
| Sex — Female, N (%) | 30 (52.6%) | 2.9 ± 5.9 | |
| Years of Education, Mean ± SD | 14.9 ± 2.7 | Rho= -0.365 | **0.005**^ |
| Race — White, N (%) | 53 (93.0%) | 2.8 ± 6.0 | 0.187# |
| Race — Non-White, N (%) | 4 (7.0%) | 5.2 ± 6.2 | |
| Area Deprivation Index (ADI), Mean ± SD | 4.7 ± 2.6 | Rho = 0.313 | **0.020**^ |
| ADI National Percentile, Mean ± SD | 52.1 ± 24.6 | Rho= 0.159 | 0.247^ |
| **Epilepsy Characteristics** |  |  |  |
| Drug-Resistant Epilepsy, Yes, N (%) | 22 (40.0%) | 3.7 ± 5.9 | 0.248# |
| Drug-Resistant Epilepsy, No, N (%) | 33 (60.0%) | 2.6 ± 6.3 | |

| **Table 1: Characteristics** | **Total Study Sample (N = 57)** | **FAQ Scores (Average ± SD)** | **p-value** |
| --- | --- | --- | --- |
| Early-Onset Epilepsy (<60 years), N (%) | 36 (63.2%) | 3.9 ± 6.6 | **0.032#** |
| Late-Onset Epilepsy (≥60 years), N (%) | 21 (36.8%) | 1.5 ± 4.6 |  |
| Number of ASMs, Monotherapy or none, N (%) | 30 (52.6%) | 2.3 ± 6.6 | **0.027#** |
| Number of ASMs, Polytherapy, N (%) | 27 (47.4%) | 3.7 ± 5.3 |  |
| Active Epilepsy Duration, <10 years, N (%) | 25 (53.2%) | 1.6 ± 4.3 | 0.196# |
| Active Epilepsy Duration, ≥10 years, N (%) | 22 (46.8%) | 3.5 ± 7.0 |  |
| Localization, Frontal, N (%) | 20 (35.7%) | 1.8 ± 4.9 | **0.018%** |
| Localization, Temporal, N (%) | 32 (57.1%) | 3.0 ± 5.1 |  |
| Localization, Other (multifocal, generalized, or parieto-occipital), N (%) | 4 (7.1%) | 10.0 ± 13.4 |  |
| Focal Aware Seizures with no other co-occurring type, N (%) | 4 (7.0%) | 1.2 ± 1.9 | 0.852# |
| Other seizure type(s) present i.e, Focal with Impaired awareness of bilateral tonic-clonic seizures, N (%) | 53 (93.0%) | 3.1 ± 6.2 |  |

^: Spearman correlation #: Wilcoxon rank-sum test (two-group comparisons) %: Kruskal-Wallis test
SD: Standard Deviation, ASM: antiseizure medication; ADI: Area Deprivation Index. Bold p-values indicate $p < 0.05$ .

In terms of epilepsy characteristics, 36 (63.2%) had EOE (onset <60 years, mean age= 34.1 years) and 21 (36.8%) had LOUE (mean age =67.6 years). Temporal localization was present in 57.1% of participants, frontal in 35.7%, and other in 7.1%. The mean number of ASMs was 1.7 ± 0.9, with 47.4% on polytherapy.

### Predictors of Functional Impairment

In univariate analyses (**Table 1**), fewer years of education (Spearman rho= −0.365, p=0.005), higher ADI decile(Spearman rho=0.313, p=0.020), EOE (Mean-FAQ EOE=3.9 ± 6.6 vs LOE=1.5 ± 4.6, p=0.032), and ASM polytherapy (Mean-FAQ polytherapy=3.7 ± 5.3 vs monotherapy or no ASM=2.3 ± 6.6, p=0.027) were significantly associated with a higher/worse FAQ total score. Seizure localization was also significantly associated with FAQ scores (p=0.018). Age, DRE status, active epilepsy duration, race, and sex were not significantly associated with FAQ total score in univariate analyses.

In the multivariable logistic regression, three predictors independently and significantly predicted FAQ-defined functional impairment (FAQ ≥2) comparable to MCI (**Table 2**). EOE was associated with 7.1-fold higher odds of functional impairment compared with LOUE (OR 7.14, 95% CI 1.16–59.97, p = 0.046). Temporal lobe seizure localization was also independently associated with 4.5 times higher odds of impairment relative to frontal/other lobar localization (OR 4.46, 95% CI: 1.09–21.83, p = 0.047). Years of education was the only protective predictor, with each additional year associated with 30% lower odds of functional impairment (OR 0.70, 95% CI: 0.49–0.93, p = 0.025). The remaining predictors, age, number of ASMs, and ADI, were not associated with FAQ impairment after adjustment for covariates in the model.

**Table 2.** Multivariable Logistic Regression: Predictors of FAQ-Defined Functional Impairment (FAQ ≥2) in Older Adults with Epilepsy

| **Predictor** | **OR** | **95% CI Low** | **95% CI High** | **p-value** |
| --- | --- | --- | --- | --- |
| Age (per year) | 1.03 | 0.92 | 1.16 | 0.627 |
| Number of Anti-seizure Medications (ASMs) | 1.20 | 0.55 | 2.61 | 0.646 |
| **Early-Onset Epilepsy <60 yrs (versus LOUE ≥60)** | **7.14** | **1.16** | **59.97** | **0.046** |
| **Localization — Temporal (versus Frontal/Other)** | **4.45** | **1.09** | **21.83** | **0.047** |
| **Years of Education** | **0.70** | **0.49** | **0.93** | **0.025** |
| Area Deprivation Index | 1.11 | 0.84 | 1.47 | 0.462 |

In sensitivity analysis, education was the only factor associated with more advanced baseline functional impairment comparable to dementia(FAQ ≥5). Each additional year of education was associated with 45% lower odds of advanced dementia-level functional impairment (OR=0.55, 95% CI=0.29-0.84, p=0.021).

### Neuropsychological and Quality-of-Life Correlates of Functional Impairment

**Table 3** shows comparisons of neuropsychological and quality-of-life measures between functionally impaired (FAQ ≥2) and unimpaired (FAQ < 2) participants. WMS logical Memory II scores were significantly lower among those with functional impairment (49.9 vs 57.1, adjusted-p=0.041). MoCA total publisher score (23.1 ± 2.6 vs 25.9 ± 2.8; adjusted-p<0.001) and z-score (adjusted-p<0.001) were significantly lower among individuals with functional impairment. Among MoCA subscores, visuospatial/executive function (3.7±1.0 vs 4.5± 0.7; adjusted-p=0.022), attention (5.1 ± 0.7 vs 5.7 ± 0.6; adjusted-p=0.018), and memory/delayed recall (2.0 ± 1.5 vs 3.1 ± 1.5; adjusted-p=0.041) were significantly lower in the IADL-impaired group. Overall quality of life, as measured by the QOLIE-31, was also significantly lower in IADL-impaired participants (51.4 ± 8.6 vs 57.9 ± 10.0; adjusted-p=0.041). ADAS-Cog total scores showed a trend toward higher impairment in the functionally impaired group (14.0 ± 3.4 vs 11.7 ± 4.5; adjusted p=0.085). Other neuropsychological measures and total ECog scores did not significantly discriminate functionally impaired from unimpaired participants.

**Table 3.**
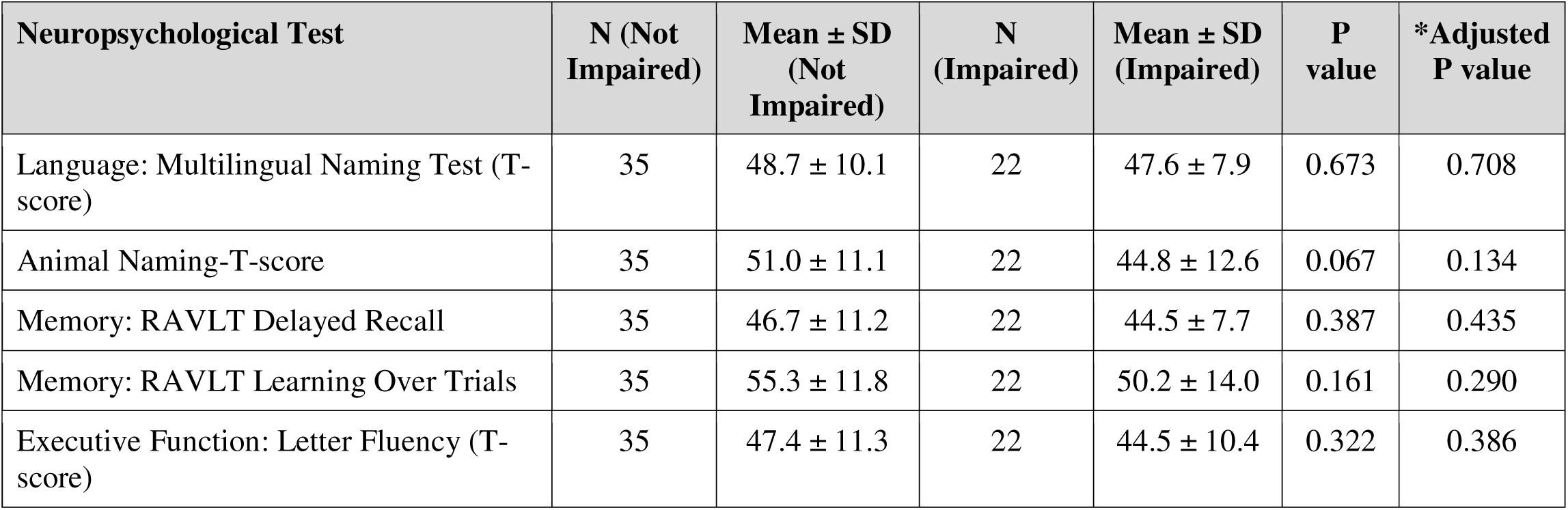
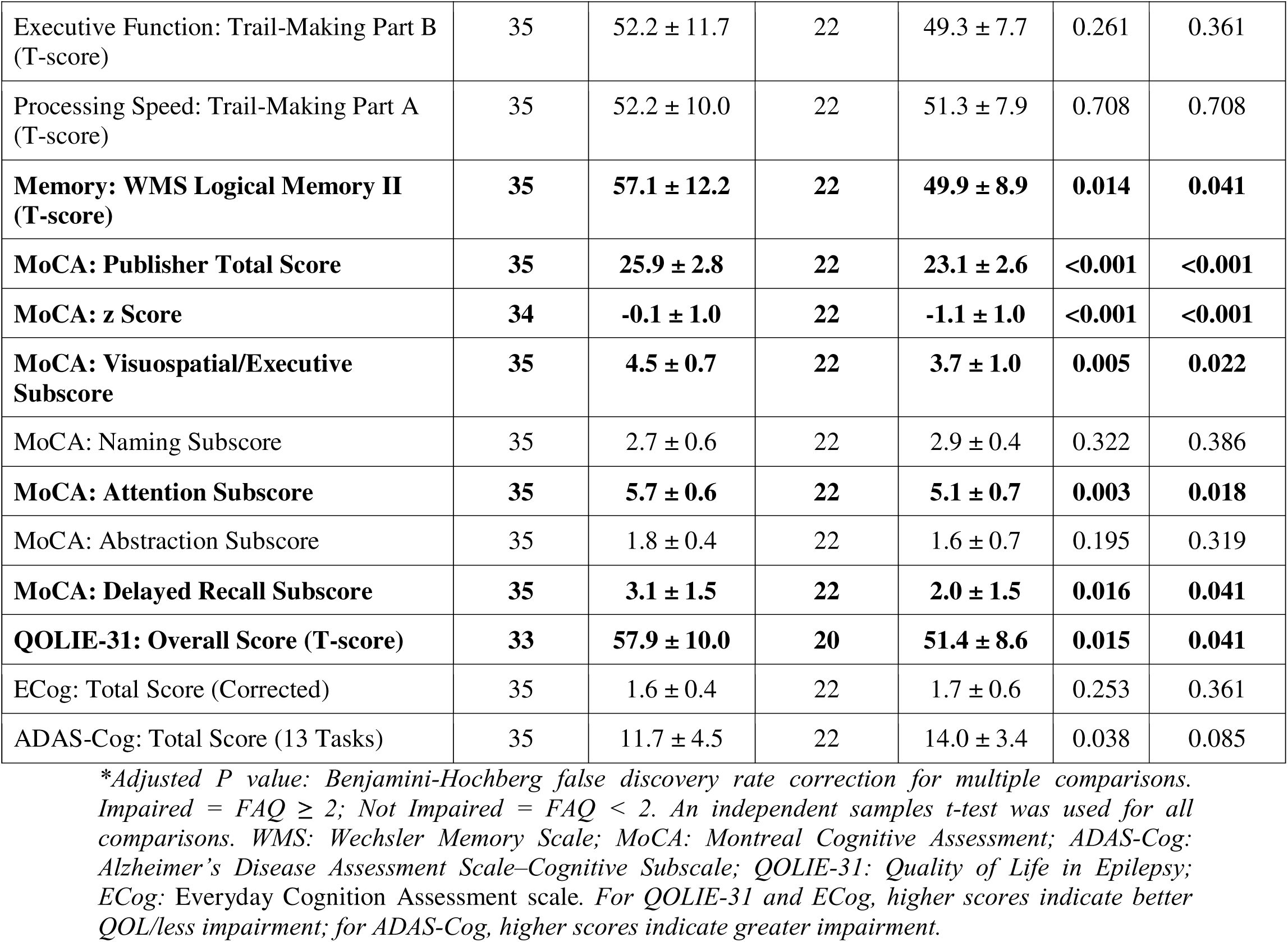
Neuropsychological and Quality-of-Life Measures by FAQ Impairment Status (Impaired = FAQ ≥ 2; Not Impaired = FAQ < 2)

**Table 4.**
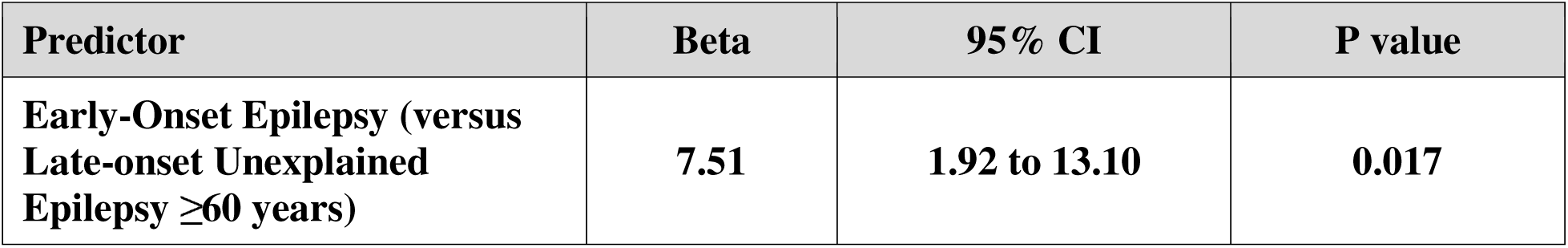
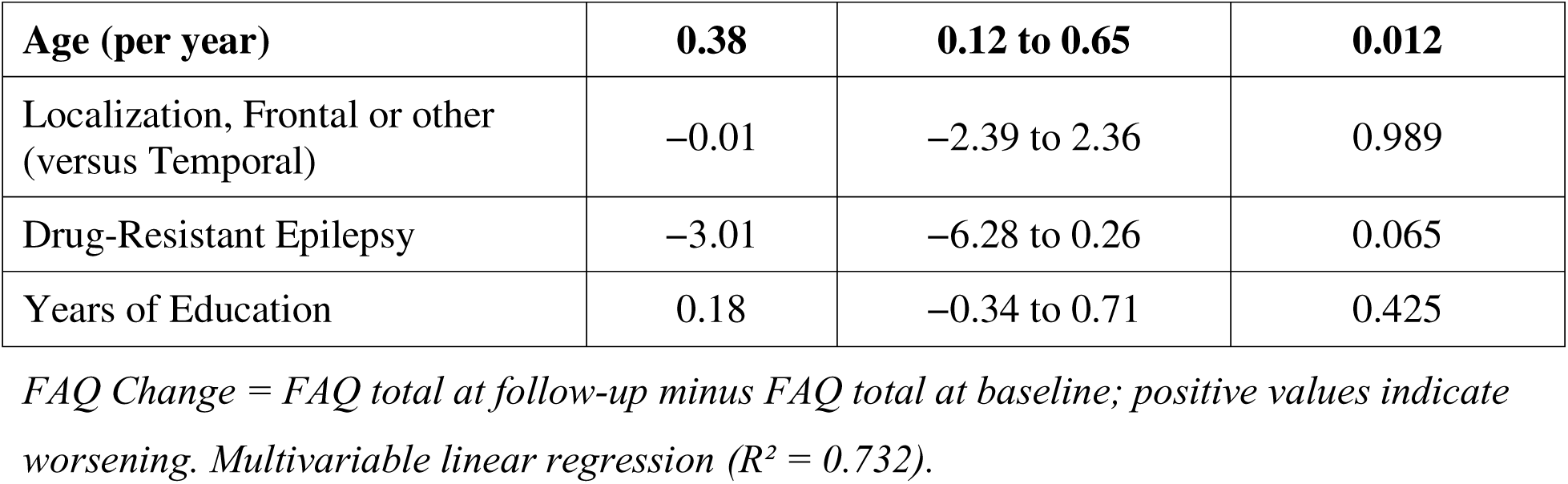
Longitudinal Analysis: Predictors of FAQ Change Score (Follow-up Minus Baseline)

### Individual Instrumental Activities of Daily Living and FAQ scores

Overall, the most commonly affected functions in which participants experienced some difficulty included maintaining a checkbook, managing bills, assembling tax records, remembering appointments, traveling independently, and playing a game of skill (**Supplemental Table 1, Supplemental Figure 1**).

Older adults with EOE demonstrated significantly higher FAQ scores on financial management items than those with LOUE (**Supplemental Table 2, Supplemental Figure 1**), specifically for the ability to manage bills and checkbook (p=0.002) and maintain tax records (p=0.006). Ability to pay bills and manage checkbook (p=0.031) was more impaired among individuals with DRE (**Supplemental Table 3, Supplemental Figure 1**) than those with non-DRE (mean 0.55 vs 0.23) and also among temporal lobe epilepsy localization (**Supplemental Table 4)** compared to frontal or other (mean temporal: 0.47 vs non-temporal: 0.18, p=0.045).

### Longitudinal Analysis: Predictors of FAQ Change

Thirteen participants had FAQ data at both baseline and a follow-up visit 2-years after enrollment. The mean change in FAQ was 0.8 ± 1.7 points over a two-year period. Individual trajectories were heterogeneous(**Figure 2**).

**Figure 2:**
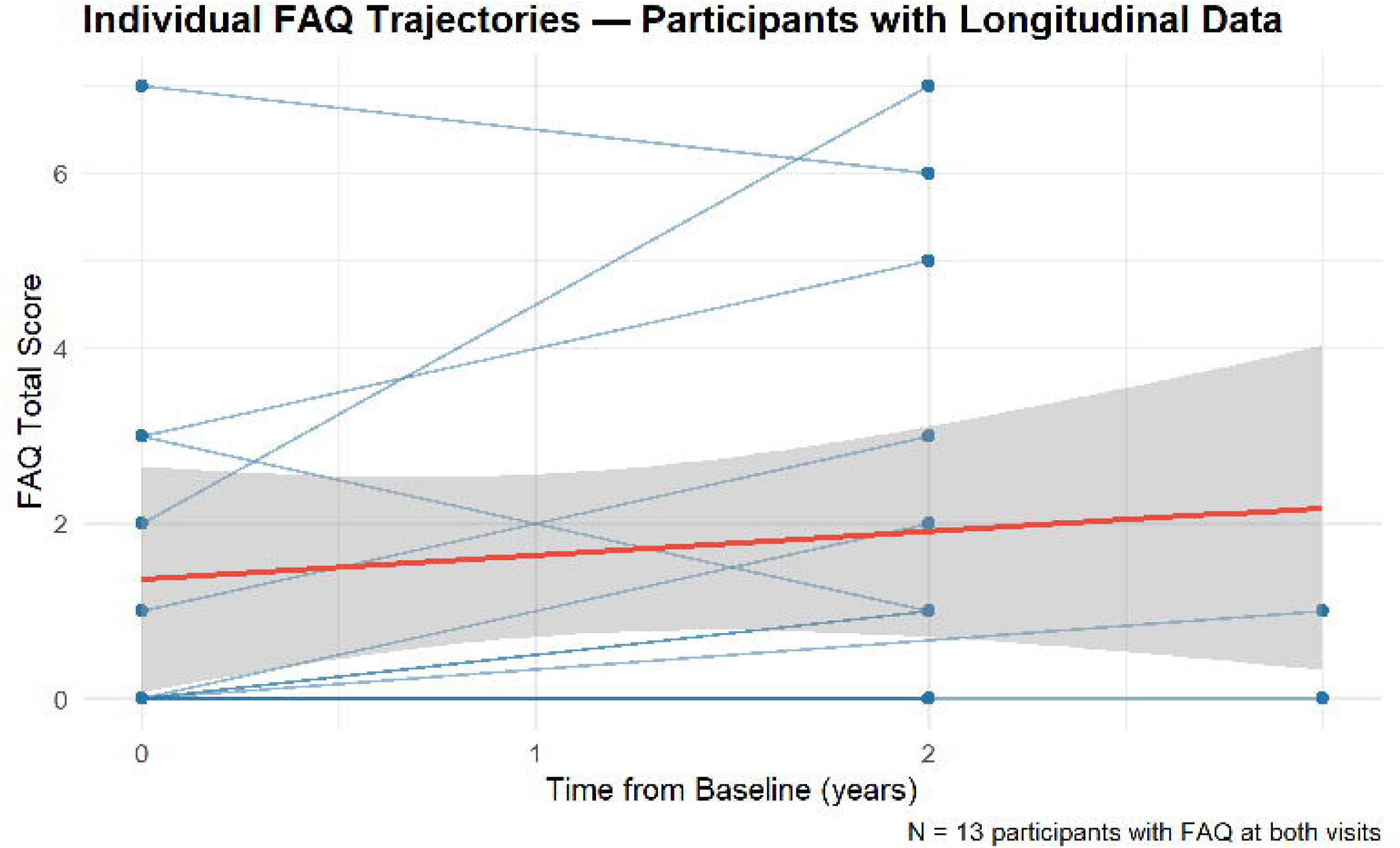
Individual FAQ (Functional Activities Questionnaire) Trajectories in Participants with Longitudinal Data.

In a multivariable (**Table 3**) linear regression model, participants with EOE had a 7.51 greater increase in FAQ score from baseline to follow-up compared to those with LOUE (Beta=7.51, 95% CI: 1.92–13.10, p=0.017). Older age was also independently associated with greater longitudinal worsening of FAQ. For each additional year of age at baseline, the FAQ score increased by 0.38 points from baseline to follow-up, after adjusting for onset group, seizure localization, education, and DRE status (Beta=0.38, 95% CI: 0.12–0.65, p=0.012). Localization, DRE status, and education were not significant predictors of longitudinal change in FAQ in the multivariable model.

### Clinically Meaningful Decline in Functional Status

Among the 13 participants with longitudinal data, four (30.8%) transitioned to a more severe functional impairment category over follow-up. Two participants with EOE progressed from MCI-level IADL-impairment(FAQ ≥2) to dementia-level IADL-impairment(FAQ >5), while two participants with LOE progressed from no functional impairment to MCI-level IADL-impairment. Four participants (30.8%) who were functionally unimpaired at baseline remained unimpaired at follow-up.

## Discussion

In this multicenter prospective observational cohort of older adults with epilepsy, we demonstrate that functional impairment of IADLs is both common and clinically meaningful, affecting nearly two in five participants, with more advanced dementia-level functional impairment observed in one in six, even in the absence of clinical dementia in our recruited cohort. We identify epilepsy-specific determinants of baseline functional impairment, most notably temporal lobe seizure localization and early-onset of epilepsy before age 60, alongside education as a protective factor. ASM polytherapy and ADI were also associated with worse baseline function. Baseline functional impairment was closely associated with verbal memory, global cognition, particularly memory, attention, and visuospatial/executive domains, and tracked with reduced quality of life. Importantly, beyond cross-sectional associations, our study is the first to examine longitudinal predictors of functional decline in epilepsy and to uniquely incorporate epilepsy-specific variables. We found that EOE and older age predict accelerated functional decline over time.

These findings position functional vulnerability as a central and under-recognized dimension of the epilepsy phenotype in later life, one that extends beyond seizure control and traditional cognitive endpoints. Prior studies of IADLs have largely focused on neurodegenerative disease or non-epilepsy-specific cohorts.^20–22^ Aside from our recent work in the ADRC cohort, demonstrating an association between active seizures and worse day-to-day function, epilepsy associated IADL impairment remains largely unexplored.^9^ The current study advances the field by providing one of the first prospective, epilepsy-focused characterizations of functional impairment and delineating disease-specific drivers of real-world functional independence.

Several aspects of our findings refine the current understanding of aging with epilepsy. We found temporal lobe localization to be associated with 4.5 times higher odds of functional impairment. This strong association suggests that temporal localization has potent effects on function. The temporal lobe underpins memory consolidation, navigation, orientation, and semantic integration,^23–25^ processes that are fundamental to complex IADLs, such as financial management, medication adherence, and independent travel. The observed pattern of deficits, particularly in financially-mediated and prospective memory-dependent tasks, aligns with this neurobiological framework and suggests that temporal lobe dysfunction may preferentially disrupt the integration of memory and executive processes^26^ required for independent living. This extends prior epilepsy research, which has largely focused on cognitive test performance,^27^ by directly linking localization to functional outcomes.

Education emerged as the only consistently protective demographic factor, with each additional year of education associated with 30% lower odds of IADL impairment and 45% lower odds of more advanced dementia-level IADL impairment. This finding supports the concept of a cognitive reserve^28^ framework in epilepsy,^29,30^ whereby higher education may mitigate the functional consequences of network disruption through compensatory cognitive strategies or more efficient neural processing. Notably, this effect persisted after accounting for epilepsy characteristics, suggesting that reserve operates independently of disease burden. In contrast, traditional markers of epilepsy severity, including drug resistance and seizure type, were not associated with baseline functional impairment. This dissociation challenges a seizure-centric view of disability and instead highlights the importance of cognitive reserve and network resilience in shaping functional outcomes. ASM polytherapy was associated with worse functional status in univariate analysis (although not significant after multivariable adjustment). This is consistent with evidence that polytherapy carries a greater burden of neurocognitive adverse effects than monotherapy. ASM polytherapy has dose-dependent effects on attention, psychomotor speed, and memory that can compound cognitive impairment, and polytherapy in particular is associated with an increased hazard of injury within one year in older adults with newly diagnosed epilepsy.^31–33^ Whether polytherapy contributes directly to functional decline or instead reflects more refractory, severe epilepsy warrants further study. Together, these findings suggest that education may represent a generalizable protective factor for functional independence in epilepsy, though the mechanisms, including reserve, compensatory neural recruitment, or health literacy, warrant further investigation.

Beyond individual education, a higher ADI, a measure of neighborhood socioeconomic disadvantage, was associated with worse baseline function in univariate analysis. Neighborhood disadvantage has increasingly been recognized as a determinant of cognitive aging: higher ADI values are associated with faster cognitive decline, and neighborhood disadvantage is associated with poorer cognitive function across multiple domains among middle-aged and older adults.^12,34^ ADI has been proposed as a surrogate of resilience, capturing multifactorial socioeconomic contributors to dementia risk that may operate through cognitive reserve.^35^ Although ADI did not remain significant after multivariable adjustment, its univariable association suggests that social determinants of health may shape functional vulnerability in older PWE.

MoCA, a measure of global cognition, was a robust discriminator of functional impairment. MoCA may be particularly sensitive to functional impairment because, like IADLs, it reflects the integration of multiple cognitive abilities rather than isolated cognitive measures.^36^ Individual cognitive tests may therefore be less suited to capturing the threshold at which accumulating cognitive inefficiencies translate into everyday functional difficulty; not because domain-specific assessment lacks value, but because functional independence depends on the interaction of abilities rather than any single domain. Consistent with this, the prominence of visuospatial/executive function, attention, and memory (verbal/delayed recall) suggests that IADL impairment reflects distributed network dysfunction rather than isolated deficits. These findings underscore the clinical utility of brief global cognitive screening tools in identifying PWE at risk of functional decline, while comprehensive neuropsychological evaluation remains essential for characterizing the specific cognitive profile underlying that risk and guiding targeted intervention.

The close relationship between functional impairment and reduced QOL highlights the bidirectional and clinically meaningful interplay between independence and well-being.^37^ Functional limitations likely constrain autonomy and social participation, while behavioral adaptations to epilepsy, such as fear of seizures or avoidance of complex tasks, may further exacerbate functional decline.^38^ Clinically, this finding supports the routine co-assessment of functional status and quality-of-life measures in epilepsy clinics, particularly for older patients.

At the level of individual functional domains, the most frequently affected activities included paying bills, assembling tax records, remembering appointments, traveling independently, and playing games of skill, a pattern largely concordant with our prior findings in the ADRC cohort.^9^ These tasks rely heavily on working memory, executive planning, task monitoring, and hippocampal-dependent episodic memory, the cognitive processes most consistently impaired in older PWE, with travel impairment further compounded by seizure-related driving restrictions.^9^ Epilepsy-specific factors, including longer disease duration, drug resistance, and temporal lobe localization, were each independently associated with difficulty in financial management, particularly the ability to pay bills, suggesting that cumulative seizure burden, recurrence of seizures, and temporal lobe dysfunction may preferentially disrupt the cognitive scaffolding required for complex financial IADLs.

Finally, we found that EOE was not only associated with 7 times higher odds of baseline functional impairment but also predicted accelerated worsening of function over time compared with LOUE in longitudinal follow-up, independent of age, localization, DRE status, and education. This is notable as LOUE is often presumed to confer greater cognitive risk due to its association with neurodegenerative processes.^1,39^ Instead, our results suggest that EOE through cumulative disease burden, reflecting decades of seizure exposure, chronic ASM use, and long-standing network reorganization, may be a key driver of accelerated functional decline and functional aging in epilepsy.^40,41^ Older age independently predicted accelerated worsening of functional outcomes, consistent with the effect of aging on epilepsy-related functional decline, supporting a model in which aging-related processes interact with epilepsy-associated network disruption to accelerate loss of independence.^42^ Although exploratory, these findings represent, to our knowledge, the first identification of epilepsy-specific longitudinal predictors of functional decline and warrant validation in larger cohorts.

While some studies have suggested a 3–5-point worsening of FAQ in MCI or dementia to be a minimal clinically important difference (MCID)^21^, an established MCID for FAQ is lacking^43^. In our cohort, nearly one-third of participants transitioned to a more severe functional impairment category over the 2-year follow-up period, suggesting that meaningful changes in day-to-day independence occurred in a substantial subset. Prior studies in AD cohorts have reported average biannual FAQ worsening of 1.23 points in MCI and 1.77 points in dementia, with minimal change among cognitively normal individuals.^22^ In comparison, the mean worsening of 0.8 points observed in our cohort over a 2-year period appears modest at the group level. However, reliance on mean change alone may underestimate clinically relevant decline, as functional trajectories were heterogeneous and several participants crossed clinically recognizable thresholds of impairment. Emerging evidence further suggests that the clinical importance of FAQ change may depend on baseline cognitive status, with smaller changes potentially carrying greater significance among individuals with normal cognition or MCI than among those with established dementia.^20^

### Limitations

Several limitations warrant consideration. The baseline sample size of 57, with only 13 participants with longitudinal data, limits statistical power. Therefore, our results require validation in larger longitudinal cohorts. Additionally, cross-sectional analyses preclude causal inference, and residual confounding by unmeasured factors, such as subclinical neurodegenerative pathology, cannot be excluded. The use of an informant-based measure introduces potential reporting bias, although this approach is standard in functional assessment and may be more reliable than patient/participant self-report.^44^ Driving restrictions in individuals with active seizures may contribute to FAQ scores via the travel item,^45^ though the mandatory driving restriction is clinically relevant and intrinsic to active epilepsy. The cohort is predominantly white and drawn from tertiary academic epilepsy centers, which limits generalizability to community-dwelling or more diverse populations. Finally, the cohort excludes individuals with prior epilepsy surgery, meaning findings may not apply to surgical candidates.

## Conclusion

Functional impairment is common in older PWE and is shaped by epilepsy-specific factors, including age of epilepsy onset and localization, ASM burden, cognitive reserve, ADI, education attainment, and global cognitive performance, with EOE and older age predicting accelerated decline over time. At a broader level, these results support a shift in how outcomes in older adults with epilepsy are conceptualized. IADLs represent an integrative endpoint that reflects the convergence of cognitive, behavioral, and environmental factors, and may serve as a sensitive marker of vulnerability in older PWE. These findings suggest that functional limitations may emerge earlier and more frequently than previously appreciated, even in PWE without overt dementia. In some individuals with EOE, this may also reflect lifelong under-attainment of specific IADLs secondary to ongoing epilepsy.

Despite its limitations, this study provides one of the first epilepsy-specific and longitudinally informed characterizations of functional impairment in older PWE. Future work should also determine whether functional decline represents an early marker of neurodegenerative comorbidity, a consequence of cumulative epilepsy burden, or an interaction of both processes.

Routine incorporation of IADL assessment with the FAQ, combined with global cognitive screening and quality-of-life evaluation, is warranted across the epilepsy clinic encounter for older adults. The identification of epilepsy-specific predictors of both baseline impairment and longitudinal decline advances the field to a function-centered framework for epilepsy care, one that prioritizes independence and quality of life alongside seizure control.^46^

## Data Availability

All data produced in the present study are available upon reasonable request to the authors

## Ethics approval statement

This study received ethical approval from the Institutional Review Boards at the University of California, San Diego, the Cleveland Clinic, and the University of Wisconsin-Madison.

## Patient consent statement

All participants provided written informed consent.

## Author Contributions

IZ conceptualized the idea. IZ conducted the formal analysis. IZ wrote the original draft. IZ, AR, KA, BPH, RMB, JK, AFS, JK, MQ, AS, KJB, MW, IW, JJS, JJ, VP, LF, and CRM supervised, validated, revised, reviewed, and edited the manuscript for intellectual content.

## Acknowledgments

This research is funded by NINDS/NIA R01NS120976. AR is funded by NINDS K23NS138682. RMB is supported by R01 NS120976, NINDS R01 NS135080, NINDS U54 NS092090, and Cleveland Clinic Epilepsy Center. I.Z. is supported by the American Epilepsy Society (Award ID: 1067206), Alzheimer’s Association (Grant: AACSFD-22-974008), BAND foundation, UVA Brain Institute, and NIH/NIA (K23AG084893). C.R.M. is supported by R01 NS120976, R01 NS124585 and R01 NS122827. B.P.H. is supported by R01NS123378, R01-NS111022, R01-NS120976, and R01-NS117568.

