## Supplemental Table 1 for "Instrumental Activities of Daily Living in Older Adults with Epilepsy: A Cross-Sectional and Longitudinal Multicenter Study"

**Supplemental Table 1: FAQ Individual Item Score Distributions** (Recoded: 0=Normal, 1=Difficulty, 2=Requires Assistance, 3=Dependent)

| **Item** | **N** | **Score 0 (Normal)** | **Score 1 (Difficulty)** | **Score 2 (Assistance)** | **Score 3 (Dependent)** | **Mean** | **SD** |
| --- | --- | --- | --- | --- | --- | --- | --- |
| Q1: Bills/Checkbook | 55 | 42 | 9 | 2 | 2 | 0.35 | 0.73 |
| Q2: Tax Records | 56 | 41 | 10 | 3 | 2 | 0.39 | 0.76 |
| Q3: Shopping | 57 | 49 | 2 | 4 | 2 | 0.28 | 0.75 |
| Q4: Games/Hobby | 56 | 48 | 7 | 0 | 1 | 0.18 | 0.51 |
| Q5: Stove/Coffee | 57 | 52 | 2 | 0 | 3 | 0.19 | 0.69 |
| Q6: Balanced Meal | 57 | 47 | 5 | 2 | 3 | 0.32 | 0.78 |
| Q7: Current Events | 57 | 52 | 0 | 2 | 3 | 0.23 | 0.76 |
| Q8: TV/Book/Magazine | 57 | 51 | 2 | 2 | 2 | 0.21 | 0.67 |
| Q9: Remember Appointments | 57 | 43 | 7 | 5 | 2 | 0.40 | 0.80 |
| Q10: Travel/Transport | 57 | 41 | 10 | 1 | 5 | 0.47 | 0.91 |

**Supplemental Table 2. FAQ Item Scores: Early-Onset Epilepsy (EOE) vs Late-Onset Epilepsy (LOE)**

| **FAQ Item** | **EOE <60 (N=36) Mean (SD)** | **EOE <60 (N=36) N** | **LOE ≥60 (N=21) Mean (SD)** | **LOE ≥60 (N=21) N** | **W statistic** | **P value** |
| --- | --- | --- | --- | --- | --- | --- |
| **Q1: Bills/Checkbook** | **0.54 (0.85)** | **35** | **0.00 (0.00)** | **20** | **480.0** | **0.002** |
| **Q2: Tax Records** | **0.58 (0.87)** | **36** | **0.05 (0.22)** | **20** | **484.5** | **0.006** |
| Q3: Shopping | 0.36 (0.80) | 36 | 0.14 (0.65) | 21 | 430.5 | 0.154 |
| Q4: Games/Hobby | 0.22 (0.59) | 36 | 0.10 (0.31) | 20 | 385.0 | 0.490 |
| Q5: Stove/Coffee | 0.22 (0.72) | 36 | 0.14 (0.65) | 21 | 401.0 | 0.448 |
| Q6: Balanced Meal | 0.42 (0.84) | 36 | 0.14 (0.65) | 21 | 451.0 | 0.070 |
| Q7: Current Events | 0.28 (0.81) | 36 | 0.14 (0.65) | 21 | 401.0 | 0.448 |
| Q8: TV/Book/Magazine | 0.25 (0.69) | 36 | 0.14 (0.65) | 21 | 410.5 | 0.320 |
| Q9: Remember Appointments | 0.50 (0.81) | 36 | 0.24 (0.77) | 21 | 459.5 | 0.075 |
| Q10: Travel/Transport | 0.53 (0.97) | 36 | 0.38 (0.80) | 21 | 404.5 | 0.585 |

*Wilcoxon rank-sum test (ordinal outcome, non-normal distribution).*

**Supplemental Table 3. FAQ Item Scores: Drug-Resistant Epilepsy (DRE) vs Non-DRE**

| **FAQ Item** | **DRE (N=22) Mean (SD)** | **DRE (N=22) N** | **Non-DRE (N=33) Mean (SD)** | **Non-DRE (N=33) N** | **W statistic** | **P value** |
| --- | --- | --- | --- | --- | --- | --- |
| **Q1: Bills/Checkbook** | **0.55 (0.80)** | **22** | **0.23 (0.67)** | **31** | **431.5** | **0.031** |
| Q2: Tax Records | 0.55 (0.80) | 22 | 0.31 (0.74) | 32 | 423.0 | 0.114 |
| Q3: Shopping | 0.41 (0.80) | 22 | 0.18 (0.73) | 33 | 418.5 | 0.103 |
| Q4: Games/Hobby | 0.18 (0.39) | 22 | 0.16 (0.57) | 32 | 381.0 | 0.389 |
| Q5: Stove/Coffee | 0.23 (0.69) | 22 | 0.18 (0.73) | 33 | 388.5 | 0.389 |
| Q6: Balanced Meal | 0.41 (0.85) | 22 | 0.24 (0.75) | 33 | 400.5 | 0.323 |
| Q7: Current Events | 0.32 (0.84) | 22 | 0.18 (0.73) | 33 | 388.5 | 0.389 |
| Q8: TV/Book/Magazine | 0.14 (0.47) | 22 | 0.21 (0.74) | 33 | 361.5 | 0.973 |
| Q9: Remember Appointments | 0.36 (0.73) | 22 | 0.45 (0.87) | 33 | 347.0 | 0.727 |
| Q10: Travel/Transport | 0.55 (1.06) | 22 | 0.45 (0.83) | 33 | 359.5 | 0.949 |

*Wilcoxon rank-sum test (ordinal outcome, non-normal distribution).*

*.***Supplemental Table 4. FAQ Item Scores: Temporal vs Non-Temporal Localization**

| **FAQ Item** | **Temporal (N=32) Mean (SD)** | **Temporal (N=32) N** | **Non-Temporal (N=24) Mean (SD)** | **Non-Temporal (N=24) N** | **W statistic** | **P value** |
| --- | --- | --- | --- | --- | --- | --- |
| **Q1: Bills/Checkbook** | **0.47 (0.76)** | **32** | **0.18 (0.66)** | **22** | **437.5** | **0.045** |
| Q2: Tax Records | 0.50 (0.80) | 32 | 0.26 (0.69) | 23 | 431.5 | 0.168 |
| Q3: Shopping | 0.19 (0.59) | 32 | 0.42 (0.93) | 24 | 340.0 | 0.236 |
| Q4: Games/Hobby | 0.12 (0.34) | 32 | 0.26 (0.69) | 23 | 348.0 | 0.586 |
| Q5: Stove/Coffee | 0.16 (0.57) | 32 | 0.25 (0.85) | 24 | 386.0 | 0.960 |
| Q6: Balanced Meal | 0.31 (0.74) | 32 | 0.33 (0.87) | 24 | 390.0 | 0.891 |
| Q7: Current Events | 0.22 (0.71) | 32 | 0.25 (0.85) | 24 | 386.0 | 0.960 |
| Q8: TV/Book/Magazine | 0.09 (0.39) | 32 | 0.38 (0.92) | 24 | 342.0 | 0.201 |
| Q9: Remember Appointments | 0.38 (0.66) | 32 | 0.46 (0.98) | 24 | 398.5 | 0.760 |
| Q10: Travel/Transport | 0.56 (0.91) | 32 | 0.38 (0.92) | 24 | 453.5 | 0.150 |

*Wilcoxon rank-sum test (ordinal outcome, non-normal distribution).*

**Supplemental Figure 1:**

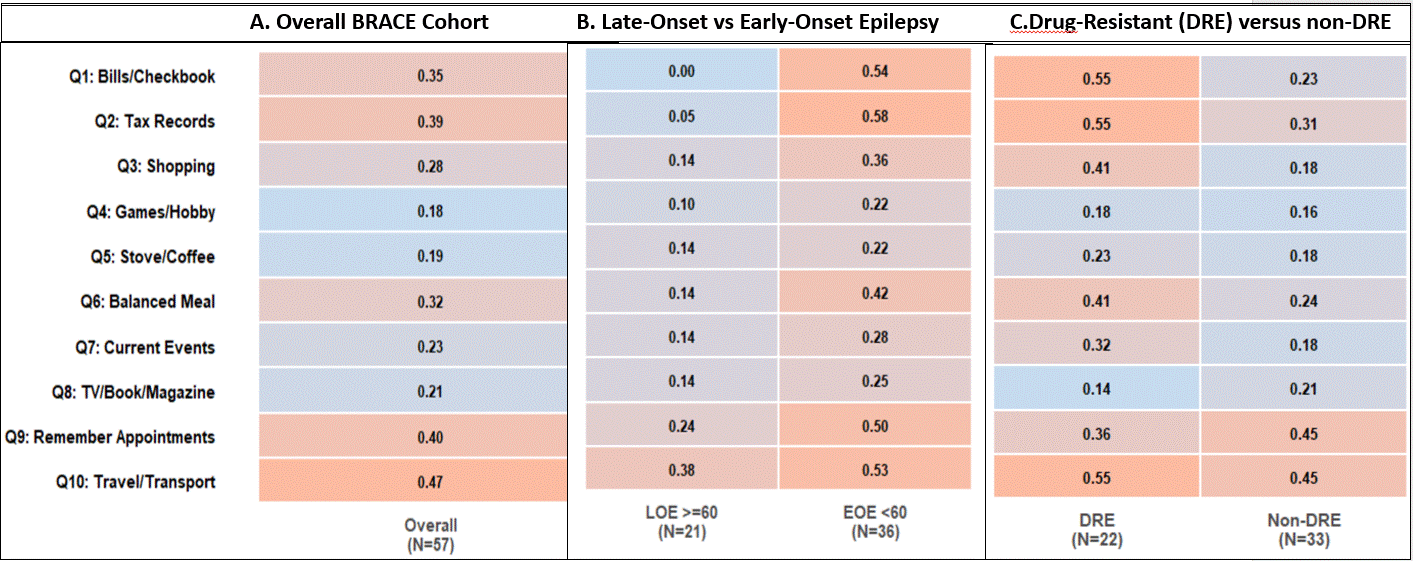
